# Co-developing a novel intervention to promote wellbeing of family caregivers of individuals with spinal cord injury: Research protocol

**DOI:** 10.1101/2024.06.09.24308666

**Authors:** Somayyeh Mohammadi, Beth Erlander, Heather Cathcart, Julie M. Robillard, David GT Whitehurst, Elena Pauly, Brooke Pagé, Sophia Sauvageau, William C. Miller

## Abstract

Family caregivers of individuals with spinal cord injury (fcSCI) are responsible for providing assistance with activities of daily living for individuals with spinal cord injury (SCI), which can include emotional support and physical assistance. Over time, providing daily support can put fcSCI at risk of experiencing caregiver burden. Burden and distress can have a substantial impact on fcSCI’s wellbeing as well as their ability to respond to the needs of the individual with SCI. A direct predictor of fcSCI burden is their appraisal of their ability to cope with the individual with SCI’s illness. Therefore, supporting fcSCI’s access to education relevant to their role and the health and wellbeing of the individual with SCI can help decrease levels of burden. The purpose of this study is to evaluate the fidelity of the intervention named COMPANION and the study protocol for an eHealth approach designed to improve outcomes for fcSCI. COMPANION, co-created with caregiver partners, is an online education program designed to provide accessible education and support for fcSCI. A concurrent mixed methods study including a feasibility randomized controlled trial will be conducted to (i) assess the process, resource, management and treatment indicators; (ii) estimate the parameters needed for a full-scale, multi-site randomized controlled trial and (iii) evaluate the effect that COMPANION has on caregiver clinical outcomes compared to a control group. The primary outcome measured will be fcSCI’s burden in addition to secondary outcomes measuring depression, anxiety, relationship satisfaction, and quality of life. The fcSCI in the intervention group will be given access to COMPANION (at T1) and data will be collected again after three months (T2) and six months (T3) to capture the impact of COMPANION on fcSCI’s psychological wellbeing. Study results will evaluate whether the full study can and should be conducted and will lead to refinement of COMPANION.

## Introduction

Approximately 360,000 individuals live with spinal cord injury (SCI) in the United States (1) and a further 86,000 in Canada (2). An SCI causes a substantial disruption in one’s life habits, routines and level of independence. In many cases, individuals with SCI need considerable human assistance to perform any activities that exceed their motor and sensory capability (3). Family caregivers, defined as “any relative, partner, friend or neighbor who has a significant personal relationship with, and provides a broad range of assistance for, an older person or an adult with a chronic or disabling condition” (4, p1) of individuals with SCI (fcSCI) often provide the majority of physical care to their family members with SCI (5). Furthermore, individuals with SCI may experience higher levels of depression and anxiety and lower levels of life satisfaction than the general population (6), therefore, fcSCI may also provide emotional support alongside physical support. Providing physical and emotional care and support can have a substantial impact on fcSCI’s physical and psychological wellbeing (5). Many fcSCI report physical, emotional and financial challenges associated with their caregiving roles and tend to neglect their own mental and physical health (7–9). Over time, these experiences put fcSCI at high risk of burnout from increased caregiver burden, which is associated with decreased wellbeing (5,8).

Caregiver burden is defined as “the extent to which caregivers perceive that caregiving has had an adverse effect on their emotional, social, financial, physical or spiritual functioning” (10, p437). Previous research indicates that 40-46% of fcSCI report moderate to severe levels of caregiver burden, due to the long-term nature of the injury and the caregiver role (10). The burden associated with caregiving also influences fcSCI’s ability to respond to the needs of the individual with SCI, and the long-term impact of caregiving demands on fcSCI may result in decreased support and quality of care for the individual with SCI. Hence, attending to risk factors and interventions that target the well-being of fcSCI is crucial.

While family caregivers of individuals with other chronic conditions, especially dementia, receive considerable attention in research (11–13), the development and evaluation of interventions that address the unique challenges faced by fcSCI have received less attention. The informal caregiving integrative model (ICIM) (14), describes several factors that determine caregiver burnout, which can be divided into three main categories: (i) *caregivers*: caregivers’ sociodemographic characteristics, psychological factors (e.g., emotion regulation, perceived competence) and physical state; (ii) *caregiving setting*: primary stressors (e.g., living with the care-recipient, being a spouse) and secondary stressors (e.g., having a reduced social life, loss of friends); and (iii) *social environment*: informal and partner support, professional support (e.g., availability of professional support and relationship with the health care providers), and sociocultural environment (e.g., how caregiving role is perceived in the society). While some of the factors impacting caregiver burden cannot be modified (e.g., gender or relationship), other factors such as emotion regulation or relationships with health care providers are modifiable. Specifically, the ICIM (14) explains that fcSCI’s appraisal of their ability to cope with the individual with SCI’s illness is a direct predictor of their burden. FcSCI’s coping appraisal is directly associated with their need for available resources and knowledge regarding the individual with SCI’s health condition. Therefore, helping fcSCI access education can increase their appraisal of their coping ability and lower levels of burden.

A recent scoping review by McKay and colleagues (15) suggests that fcSCI maintain better wellbeing if they have enough leisure activities, strong problem-solving skills, and practical and emotional support (15). In a recent qualitative study of Canadian fcSCI, Jeyathevan and colleagues (16) identified that access to community and social support, a better relationship with the individual with SCI, and mastery in caregiving could help fcSCI provide sustainable care for individuals with SCI and for themselves. While the findings of Jeyathevan and colleagues (16) identified several critical needs of fcSCI, few studies have investigated the impact of different types of interventions on family caregivers’ outcomes—the studies that have been conducted have focused mainly on teaching problem-solving skills (16–20). Additionally, even though rehabilitation facilities invite fcSCI to join educational sessions, these sessions primarily focus on the patients’ needs. Compounding this challenge, fcSCI often have limited time, making attendance at in-person sessions difficult.

In our objective to improve outcomes for fcSCI, this study will evaluate the implementation fidelity of an innovative program for health education with online approaches (eHealth) intervention that aims to improve the wellbeing of fcSCI. COMPANION, cocreated with caregiver partners and health care professionals, is an interactive eHealth education program that is based on adult learning principles (21) and a user-centered approach (22). The goal of COMPANION is to address fcSCI’s specific challenges in caring for themselves and for the individuals with SCI and facilitate the provision of accessible online education. We envision COMPANION will improve access to existing knowledge for fcSCI and individuals with SCI and provide customized (e.g., developed through understanding caregivers’ perspective), interactive (e.g., through quizzes) and engaging eHealth education (e.g., new educational videos and audios) in the form of online modules, which are essential aspects of an eHealth tool (23) to better prepare and support fcSCI for providing care for themselves and the individuals with SCI.

## Materials and methods

This paper provides the details of a study protocol for a concurrent mixed methods study, including a feasibility randomized controlled trial (RCT). The purpose of the feasibility RCT is to (i) assess the fidelity of the process, resource, management and treatment indicators and (ii) estimate the parameters needed for a full-scale, multi-site randomized controlled trial (24). Secondary objectives will be to assess the effect of COMPANION on primary and secondary clinical outcomes for fcSCI (24,25).

### Study design

To assess the feasibility and intervention fidelity of COMPANION, a six-month feasibility RCT with a 1:1 allocation ratio will be used, comparing COMPANION to the standard education and resources available to fcSCI. The study incorporates quantitative and qualitative design methods administered through surveys and a final exit interview. In conjunction with RCT studies in rehabilitation the use of qualitative methods has been recommended to spot unexpected variables and findings (26,27). The interview will be an opportunity for participants to share perspectives about their experience using COMPANION, with findings used to interpret the clinical outcomes and, when integrated with quantitative findings, identify patterns and paradoxes between the results.

This research has been funded by the Craig H. Neilsen Foundation PSR Pilot Grant (PSR2-17), Award Number: 865706. The research is sponsored by the University of British Columbia and has been approved by the University of British Columbia Behavioural Research Ethics Board. This trial has been registered on ClinicalTrials.gov with ID NCT06364813.

### Sample size and inclusion and exclusion criteria

Suggested sample sizes for a feasibility RCT range from 12-30 per experimental group (28,29). We aim to recruit 20 participants per group. Inclusion criteria for participants will be, at the time of recruitment: (i) family caregiver of an adult individual with SCI (similar to previous studies of this population (30,31); in this study, family caregiver is defined as an individual who is primarily responsible for providing immediate informal daily care for a relative with SCI), (ii) >18 years old, (iii) live with the individual with SCI in the community, (iv) live in North America, and (v) the spinal cord injury of the individual with SCI has not happened within the previous six months. FcSCI with major medical and physical conditions that require routine visits to medical doctors (e.g., cancer) will be excluded from the study. FcSCI will be excluded if they provide care to individuals with SCI who are still patients in a rehabilitation facility.

### Study setting and recruitment

FcSCI will be approached to participate: (i) indirectly by sending letter and email invitations to previous patients or research participants of our center (GF Strong Rehabilitation Center and the International Collaboration on Repair Discoveries (ICORD)) and asking them to invite their family caregiver to contact us, and (ii) directly by asking our caregiver partners, Wives and Girlfriends of Spinal Cord Injury (WAGS of SCI), to approach their members (approximately 6,000 members, mainly in North America) and invite them to participate in this study. We will use social media advertisements to facilitate recruitment. Participants will be recruited internationally from locations across North America. To be inclusive of participation of individuals from remote and rural areas, all participants will be invited to participate digitally through a virtual meeting platform. In-person participation will not be offered for study participants regardless of physical proximity to the research team. After screening to ensure participants meet the eligibility criteria, all participants will complete written, informed consent.

### Randomization and masking

Upon successful enrollment and baseline data collection, participants will be randomized by research staff using the online service Sealed Envelope (32) with a block size that will be undisclosed to the study manager. The research staff is responsible for randomization and the delivery of the intervention while the study manager, who is responsible for enrolling participants and administering data collection, will be masked to the participants’ group allocation. To mitigate performance bias, participants will be instructed not to discuss their program or group allocation to any member of the research team except the research staff, and both the study manager and research staff will reinforce this point when contacting participants. The research staff will be responsible for receiving emails from participants and removing information related to group allocation before forwarding participant emails to the study manager.

### Intervention

COMPANION was designed through a collaboration between researchers, healthcare providers, and family caregivers (end users and stakeholders) using the Technology Co-Design Model (33,34). The current preliminary version of COMPANION consists of eight self-paced online modules, which can be accessed remotely from any location.

### Intervention Group

Participants in the intervention group will be able to choose the order in which they complete the COMPANION modules and will be encouraged to complete them all. Participants in the intervention group will be encouraged to invite the individual with SCI to view the COMPANION materials with them as several of the modules include exercises that the participants can do with the individual with SCI and information regarding processes that the fcSCI navigates with the individual with SCI (such as changes in their relationship, considerations for hiring a home care aide and accessing financial and legal support). Participants will receive the web address, instructions to access the website using personalized encrypted login information, and a guideline on how to use the program. Modules have been designed so they can be stopped at any point, with progress automatically saved. Module formats include embedded video clips, voiceovers, and additional resources including links to other available information. As topics are introduced in the course, participants will be reminded that the content in the modules is not a substitution for expert professional advice and will be instructed to consult with their healthcare providers when needed. If participants have questions regarding the content of the modules, they can submit their questions to the research staff. Research staff will review the submitted questions and if necessary forward them to one of the experts on the expert panel, consisting of a selected team of professionals who have experience working with individuals with SCI and their family caregivers. The selected expert will provide additional resources or information to address that participant’s question. The experts on the proposed project will not provide personal interventions to family caregivers; instead, they will focus on providing information to address the submitted questions only. Questions submitted by participants during the feasibility study will be used to modify and improve COMPANION. The research team will record participants’ submitted questions and the amount of time spent addressing each question and responding to participants. This information will also be used to improve further and modify COMPANION.

In the first three months of the feasibility RCT, research staff will remotely monitor online analytics (e.g., login frequency, module progression) and will contact a participant if no online activity is noted in a two-week period to promote use and troubleshoot potential technical problems.

### Control group

Control group participants will receive the usual care available at their local rehabilitation clinic. Immediately upon reaching the 6-month time point, control group participants will be given access to the current version of COMPANION.

### Study timeline

The following three data collection timepoints will be used in the feasibility RCT: the time immediately following successful screening of the participant, during which baseline data will be collected, and access to COMPANION will be provided to participants in the intervention group (T1); three months after baseline data collection (T2); and six months after baseline data collection (T3).

As of the time of the submission of this publication, the study is not yet active. Neither recruitment nor data collection has begun. All data collection will be done via a secure online survey tool available through our institute (Qualtrics). After successful screening, randomization, and baseline data collection has occurred, participants will be asked to complete the T1 survey. Follow-up data will be collected at three months (T2) and six months (T3). At T1-T3, participants will respond to measures that assess primary (subjective burden) and secondary outcomes (objective burden, relationship quality satisfaction, distress, physical and mental health, and caregiving competence). The purpose of T1 is to collect baseline information on the outcomes of the study. The purpose of T2 is to capture the influence of the intervention on our psychological outcomes of interest (such as depression, anxiety, stress, and competence). T3 reflects the optimal change period for self-reporting of physical outcomes (such as physical health and functioning). Participants will receive a $50 token of appreciation for completing each timepoint.

### Feasibility indicators

The RCT’s primary feasibility indicators will be used to address the COMPANION intervention’s design and fidelity by assessing process issues, resource issues, management issues, and treatment issues. See Figure 2 for further information on the feasibility indicators.

**Figure 1.**
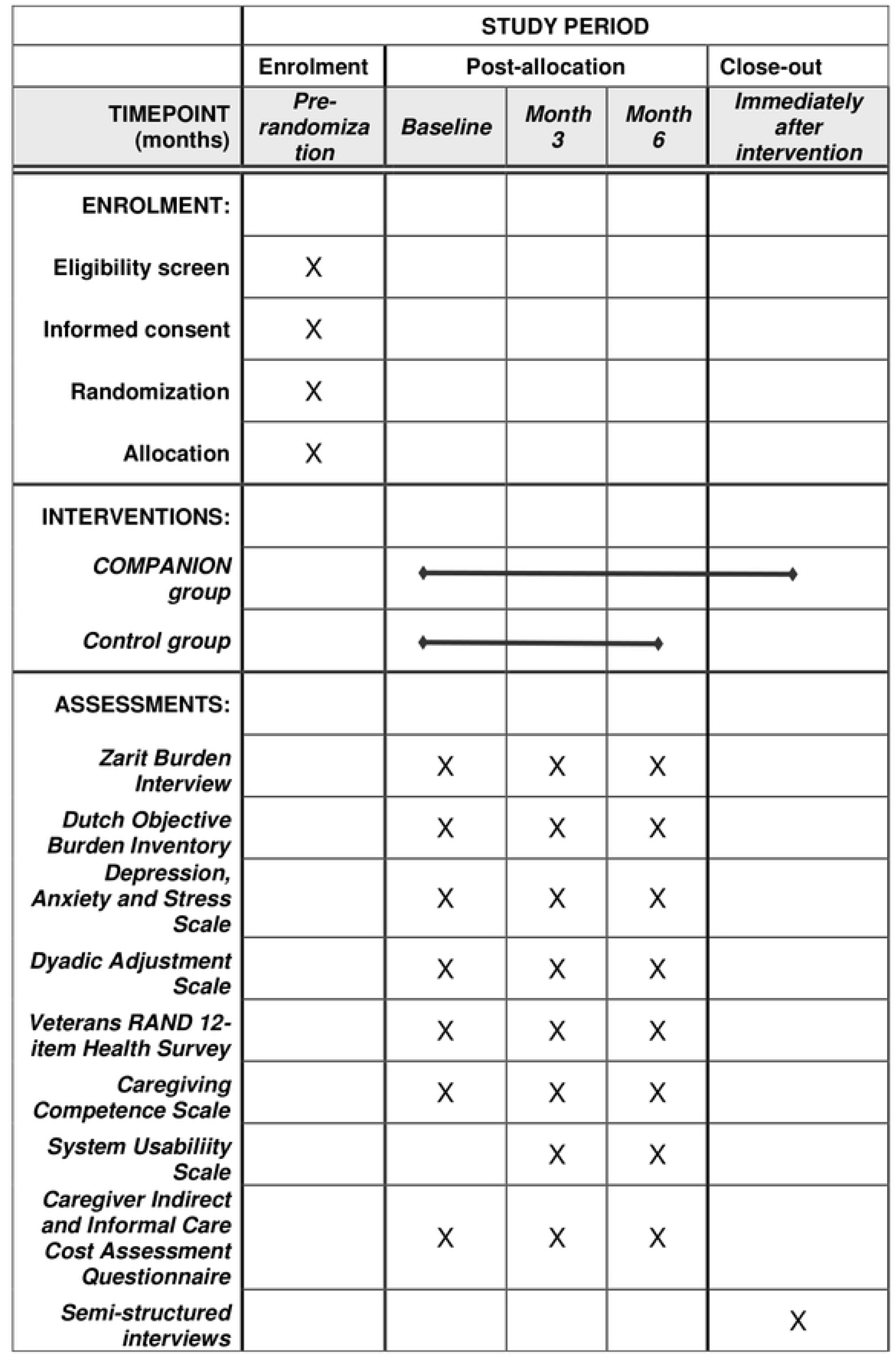
SPIRIT schedule of enrollment, interventions and assessments

**Fig 2.**
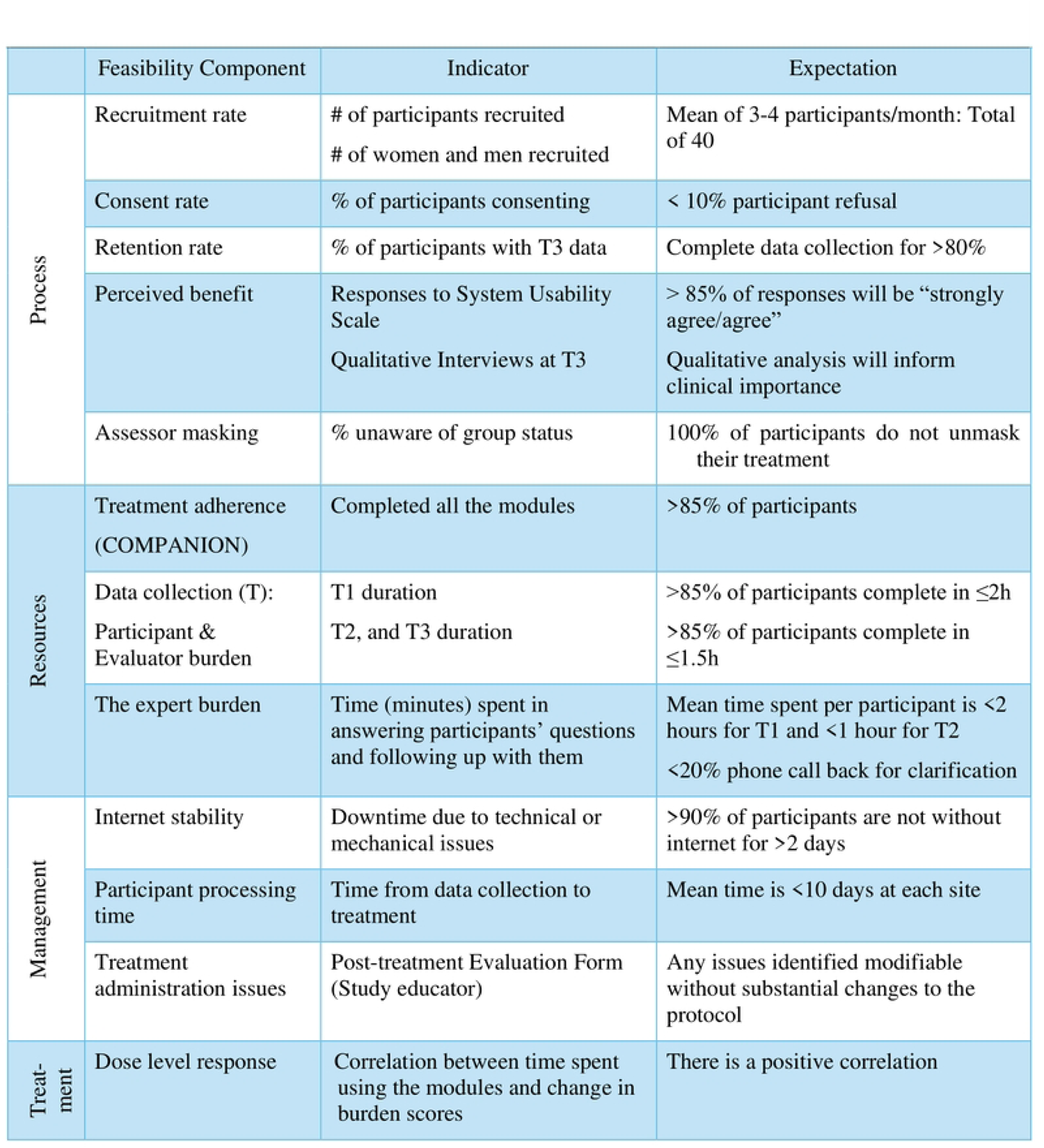
Feasibility Indicators

### Clinical outcome measures – primary outcome

Clinical outcomes will be measured via a digital survey administered at baseline, T2, and T3. The components of the survey and outcome measures are detailed below.

#### Subjective burden

The primary clinical outcome will be measured through the survey using the Zarit Burden Interview (35). Studies have demonstrated a direct association between burden and quality of life for family caregivers (19). As family caregivers’ appraisal of their ability to cope with their family member’s health condition is a direct predictor of their burden (14), and COMPANION aims to provide the required education and information for fcSCI so that they perceive and experience less challenges in providing care, we have selected fcSCI’s subjective burden as our primary clinical outcome. The short 12-item version of the Zarit Burden Interview will be used to assess subjective burden (35). FcSCI are asked to use a 5-point scale (from 0 = never to 4 = nearly always) to evaluate how often they experience certain feelings. Total scores can range from a minimum of 0 to a maximum of 48. A total score ranging between 0-10 indicates no to mild burden, 10-20 indicates mild to moderate burden, and >20 indicates high burden. This measure has been considered as valid and reliable (Cronbach’s alpha = 0.78) among fcSCI (36).

### Clinical outcome measures – secondary outcomes

The survey administered at baseline will collect information on participants’ age, sex, gender, social support, annual household income, living situation, and chronic health. FcSCI will be asked to answer similar questions for the individual with SCI. In addition to demographic information, several secondary outcomes measures will be administered at multiple timepoints. The components of the secondary outcomes measures and their timepoints are detailed below.

#### Objective Burden

The survey will be used to administer several secondary clinical outcome measures. The first secondary outcome will be measured using the 38-item Dutch Objective Burden Inventory assessing fcSCI’s objective burden (37). Each questionnaire item lists a specific caregiving activity that corresponds to one of the following four domains: personal care (e.g., helping with eating and drinking), practical support (e.g., buying groceries), motivational support (e.g., motivating to quit or reduce smoking), and emotional support (e.g., showing understanding). FcSCI are asked to rate their perceived burden for each task over the past 3 months using a 3-point scale (1 = not at all burdensome, 2 = somewhat burdensome, 3 = very burdensome). Total scores are calculated as the average of all individual answer scores, ranging from 1 to 3. A higher total score indicates higher objective burden. Testing of the measure has affirmed adequate validity and reliability among Dutch and Canadian caregivers (38,39).

#### Distress

The second secondary outcome will be measured using a short version of the Depression, Anxiety and Stress Scale (DASS) assessing distress (40). A short version of the complete 42-item DASS, the DASS-21 contains 21 items assessing depression, anxiety and stress on 3 separate sub-scales comprised of 7 items each. Participants report the intensity of their symptoms over the last week using a four-point Likert scale (from 0 = never to 3 = always). Separate total scores ranging from a minimum of 0 to a maximum of 42 will be calculated for each sub-scale. Severity levels for depression, anxiety, and stress can be interpreted based on recommended cut-off scores, with higher scores indicating more distress. Cut-off scores for the Depression scale distinguish responses as ‘Normal’ (0–9), ‘Mild’ (10–13), ‘Moderate’ (14–20), ‘Severe’ (21–27), and ‘Extremely Severe’ (28+). Testing of the DASS-21 among family caregivers has shown adequate reliability (coefficient alpha = 0.97) (41).

#### Relationship Quality Satisfaction

The third secondary outcome will be measured using the Dyadic Adjustment Scale (DAS-32) (42). The DAS-32 consists of 32 items measuring the level of relationship quality satisfaction among dyads. Items in the DAS-32 correspond to four dimensions or subscales for measuring relationship quality: Dyadic Consensus (consensus on matters of importance to marital functioning), Dyadic Satisfaction, Dyadic Cohesion (closeness experienced by the couple), and Affective Expression (demonstrations of affection and sex relations). Each item of the questionnaire corresponds to one of the four subscales, which consist of varying response scales including ordinal, Likert, and dichotomous scales. Scores for each subscale are calculated and added together to produce a total score ranging from 0 to 151 with higher total scores indicating less distress and high adjustment. Psychometric testing of the measure has demonstrated good reliability alphas for each subscale (.85 for Dyadic Consensus, .67 for Dyadic Cohesion, .76 for Affective Expression, and .82 for Dyadic Satisfaction) in the general population (42).

#### Health-related Quality of Life

The fourth secondary outcome will be measured using the Veterans RAND 12-item Health Survey (VR-12) assessing fcSCI’s health-related quality of life (43). The VR-12 includes 12 questions corresponding to eight domains: general health perceptions, physical functioning, role limitations due to physical problems, role limitations due to emotional problems, social functioning, bodily pain, vitality, and mental health (44). The eight domains can be summarized into separate physical (PCS) and mental (MCS) component scores. The VR-12 scoring algorithm is used to score PCS and MCS based on weights derived from a 1990 American population sample standard, resulting in variable minimum and maximum score values (45). Higher PCS and MCS scores indicate better health. Two further questions (i.e., 14 in total), which do not contribute to the component summaries, ask about physical health and emotional problems “compared to one year ago.” Health state values (also known as utility scores), reflecting the preferences of the Canadian population, can also be generated from VR-12 data (46). Though validation of the VR-12 has not been conducted in family caregiver populations, the VR-12 has demonstrated adequate convergent validity in other populations (47).

#### Competence

The fifth secondary outcome will be measured using the Caregiving Competence Scale (CCS) (48). The CCS (48) is used to measure the caregiver’s self-assessment of the adequacy of their performance in their role. The 4-item questionnaire scores responses using two 4-point scales with response options ranging from 1 = ‘not at all’ to 4 = ‘very much,’ and 1 = ‘not at all’ to 4 = ‘very’ (48). Score values range from 0 to 12 with higher scores indicating higher caregiving competence. The CCS has demonstrated adequate internal consistency (Cronbach’s alpha ranging from 0.78 to 0.82) in family caregiver research (49,50).

#### Usability

The sixth secondary outcome will be measured using the System Usability Scale (SUS) to capture the opinion of participants regarding the usability of COMPANION. Originally created by Brooke et al. (51), the SUS consists of a 10-item questionnaire using a 5-point scale where options range from ‘strongly agree’ to ‘strongly disagree.’ Raw scores range from 0-40 and are algorithmically converted into meaningful SUS scores ranging from 0-100. Higher scores indicate higher perceived usability of the program. Participants in the intervention group will complete this measure at T2 and T3.

#### Caregiver Costs

The final secondary outcome will be measured using the Caregiver Indirect and Informal Care Cost Assessment Questionnaire (CIIQ) to capture information estimating informal care costs (52). FcSCI will be asked to provide responses for 13 items with information including, but not limited to, work status, time missed at work, productivity, and physical and emotional caregiving duties. Calculations will be conducted using responses to items on the CIIQ to estimate fcSCI indirect and informal care costs. Total scores can range from as low as zero to any maximum number and report cost in dollars. Higher scores indicate greater financial impact due to informal care costs. The CIIQ will be coupled with a bespoke item asking about additional money fcSCI spend on themselves. To minimize concerns about recall, a one-week recall period will be used for the CIIQ and a three-month recall period will be used for the bespoke item.

#### Qualitative interview

When the feasibility RCT is complete, semi-structured interviews (∼45-60 minutes) will be used to explore fcSCI’s experiences in the intervention group with COMPANION. The option to take part in the semi-structured interview will be available to participants in the intervention group as soon as they complete T3 survey data collection. An interview guide has been created to capture multiple aspects of fcSCI’s experience using COMPANION. As a complimentary method to assess usability, the interviews will also be used in combination with the System Usability Scale (51) and feasibility indicators to evaluate the usability of COMPANION. Participants will be asked to share their perspective on advantages and disadvantages of COMPANION. A directed content analysis approach will be used to form initial coding categories from existing evidence and study hypotheses (53).

To promote the credibility and trustworthiness of the qualitative data, we will use several methods. Field notes will be used as a self-reflective tool. Member checking will be employed; i.e. participants will be invited at the end of each interview to review the preliminary findings and provide feedback to determine how well they resonate with them. Multiple investigators with diverse backgrounds will be involved in collaborative theme analysis and data coding as a form of triangulation (54). Interview results will be integrated with the quantitative results to provide a more in-depth assessment of benefits and user acceptability (55). Peer debriefing will be used to further support triangulation. To ensure the transferability of the qualitative data in our reports we will follow Consolidated criteria for reporting qualitative report guideline (COREQ) (56). Specifically, we will provide detailed information on the setting, participants, and the interviewers’ and research team’s background. We will also provide a detailed description of the data collection procedure, analyses of the qualitative data and our procedure in developing the codes and the themes to ensure the dependability of our method.

### Data management

Participant consent forms and all data collection will be administered online via Qualtrics. Data will be stored on Qualtrics and accessible only to research personnel who are granted access by the research staff. Participants will receive a link to the consent form via email with instructions about how to complete the form. After signing the online consent form, participants will schedule a video call meeting during which they will complete T1 data collection measures with the help of the study manager. Immediately after the participant and study manager have completed the T1 questionnaire, the study manager will leave the call and be replaced by the research staff, who will randomize the participant into the intervention or control group using the secure online software (32). The participants will be given instructions based on their group allocation and asked not to reveal their group to the study manager, who will remain masked to the randomization result. Participant IDs and group allocation will be recorded on a secure, password-protected digital tracking sheet. Only research personnel involved with the group allocation will have access to the tracking sheet, and the study manager will not be able to access any information on the sheet related to participant group allocation. At T2 and T3 timepoints, participants will be sent an email containing a unique link to the next Qualtrics questionnaire and will be asked to complete the survey on their own.

### Data analyses

#### Statistical analyses

All data analysis will be conducted by the research team independently from the study sponsor. Descriptive analyses will be used to describe our sample and consider clinical outcomes and study feasibility. The most current versions of R Statistical Software (57) and SPSS Software (58) will be used. The one-sample Kolmogorov-Smirnov test will be used to evaluate data distribution. Demographic and outcome variables will be summarized by groups using means, standard deviations, frequencies and proportions. Similarly to previous feasibility studies (59), the mean and the standard deviation calculated in the feasibility RCT will be used to estimate the effect size and the variance necessary for the full RCT. Focuses of the analyses will include investigating differences in primary and secondary effectiveness outcomes for the intervention and control groups at each time point. We will include an interaction between time and group for all models to account for the possibility of effect modification by time. Estimated marginal means will be used to conduct post hoc analyses, and pairwise comparisons of estimated marginal means will be conducted to examine specific differences between groups at each time point. Participant ID will be used as a random effect. Furthermore, descriptive statistics will be used to assess participants’ online usage data of the eHealth program (such as the amount of time participants spent accessing the modules) to evaluate feasibility indicators (dose level response, treatment adherence) related to resource and treatment issues (60).

#### Feasibility

Feasibility outcomes will be treated as binary based on the expectations described in Figure 2. Specific objectives will be evaluated as a “success,” indicating that no major adaptation of the protocol is before proceeding with a definitive RCT, or “revise,” indicating changes must be made before continuing with the definitive RCT.

#### Qualitative interviews

Interview transcripts will be examined using an inductive thematic analysis approach (61). Data will be analyzed to identify themes and subthemes. Investigators will generate initial codes individually and then work with the entire team, including our caregiver partners, to identify themes, review, and define themes and select examples to support the themes (61). The qualitative findings will be used to interpret the clinical outcomes, such as key intervention ingredients, relevant outcomes that might not be captured and explanation for potential conflicting quantitative results. Quantitative and qualitative findings for participants will be integrated as a form of triangulation and summarized to identify patterns and paradoxes between the results (62).

## Discussion

Health education using online approaches has been lauded for being interactive and enabling learners to engage over sustained periods. Benefits of eHealth can include improving the quality of care (37,63,64), enhancing communication between healthcare users and providers (65), reducing costs (66), and increasing access to evidence-based health information. As modules in COMPANION were designed specifically to address aspects of caregiving that are significant to fcSCI, COMPANION has the potential to increase fcSCI’s knowledge and health literacy around challenges that they face when providing care (e.g., emotional challenges, challenges in conducting medical tasks, challenges with isolation and accessing social support). By providing access to education and resources regarding factors that are known to mitigate burden in fcSCI, such as social support (67) and coping appraisal (14), COMPANION may be used to impact fcSCI’s overall feelings of burden, quality of life and wellbeing. Findings from conducting the feasibility RCT that evaluate the feasibility and acceptability of our study procedures can be used to inform the design of a definitive RCT assessing the effectiveness and impact of COMPANION on clinical outcomes compared to usual care pursued by fcSCI. Conducting a definitive RCT can further contribute meaningfully to family caregiver research and literature about mitigating the burden experienced by caregivers of individuals with spinal cord injury.

We anticipate that a large percentage of fcSCI who are approached to participate in our study will be female, due to our recruitment plan to ask WAGS of SCI to distribute recruitment materials to their members, who are primarily female fcSCI. Though women have made up the majority of fcSCI participants in recent studies with similar populations (5,68,69), we plan to employ recruitment methods aimed at diversifying the gender spectrum of fcSCI we approach by additionally contacting participants through our network of previous patients and research participants of our centre and distributing recruitment information through social media advertisements. Additionally, literature has suggested that eHealth approaches may attract a younger demographic of users who may be more experienced using online platforms than older populations, who may be discouraged from using the platform or participating (21). To support participants’ access and experience using COMPANION, participants will be sent written instructions about how to access and navigate COMPANION and its modules, encouraged to use COMPANION with their family member with SCI and instructed to contact the research staff with any questions about using the program. We also plan to monitor intervention group participants’ online activity during the study and reach out to participants if a time span of 2 weeks passes without any activity recorded. This communication will give participants an opportunity to voice any barriers they experience when using COMPANION and receive technological or other guiding support from the research staff (23).

Our knowledge translation plan will target fcSCI, individuals with SCI and clinicians across Canada and the United States, leveraging existing communication tools, such as: websites (e.g., health authorities in BC) and electronic and print newsletters of local and national audiences (e.g., SCIRE, ICORD, SCI BC). All team members including our caregiver partners will contribute to the presentations and manuscripts. We, along with our caregiver partners, will share plain language summaries with fcSCI via newsletters, social media and websites (e.g., http://wagsofsci.com/).

## Data Availability

No datasets were generated or analysed during the current study. All relevant data from this study will be made available upon study completion.

## Authors’ contributions

Conceptualization: Somayyeh Mohammadi, William C Miller

Funding acquisition: Somayyeh Mohammadi, William C Miller, Julie M. Robillard, David GT Whitehurst

Methodology: Somayyeh Mohammadi, William C Miller

Resources: Somayyeh Mohammadi, Beth Erlander, Heather Cathcart, Elena Pauly, Brooke Pagé

Writing – original draft: Somayyeh Mohammadi, Sophia Sauvageau

Writing – review and editing: Somayyeh Mohammadi, Heather Cathcart, Julie M. Robillard, David GT Whitehurst, William C Miller, Sophia Sauvageau

## Supporting information

**S1. SPIRIT Checklist.**

**S2. REB Approval.**

**S3. Trial Study Protocol**

## Notes

### Competing Interest Statement

The authors have declared no competing interest.

### Clinical Trial

The trial is registered on ClinicalTrials.gov. The ClinicalTrials.gov ID is NCT06364813 and the Protocol ID is H20-01461. The trial URL ishttps://clinicaltrials.gov/study/NCT06364813

### Funding Statement

Yes

### Author Declarations

This research has been approved by the University of British Columbia Behavioural Research Ethics Board with ID# H20-01461. Written consent was obtained.

